# How Short Is Safe? Duration of Dual Antiplatelet Therapy Before Aspirin Withdrawal After Percutaneous Coronary Intervention: A Dose-Response Network Meta-Analysis

**DOI:** 10.64898/2026.10.28.26366775

**Authors:** Jae-Sik Jang, Kang-Hee Kim, Jong Seon Park, Eun-Mi Lee, Moo-Yong Rhee

## Abstract

**Background:** Trials of P2Y12 inhibitor monotherapy after percutaneous coronary intervention (PCI) withdrew aspirin at fixed times, mostly against 12-month dual antiplatelet therapy (DAPT), and categorical syntheses disagree.

**Methods:** Bayesian model-based network meta-analysis of randomized trials of aspirin withdrawal with P2Y12 inhibitor continuation, with 12-month myocardial infarction (MI) and major bleeding modeled as functions of DAPT duration (Emax and spline). The prespecified estimand was the shortest duration at which the upper 95% credible limit of the odds ratio (OR) for MI versus 12-month DAPT fell below 1.15.

**Results:** Sixteen trials (17 randomized comparisons, 58,068 patients) contributed 929 MIs and 1,102 major bleeding events. The OR for MI was 1.31 (95% credible interval 0.95–2.09) at 0 days, 1.17 (0.96–1.43) at 30 days, and 1.09 (0.98–1.25) at 90 days. Under every model the point estimate exceeded 1.15 below about 3 weeks and fell below it from about 90 days; a 30-day change-point did not fit better. The credible bound was met at 158 days (Emax) and 282 days (spline). Bleeding decreased with shorter duration (OR 0.40 at 0 days, 0.62 at 90 days). Certainty was low for MI below 90 days and the estimand, moderate for bleeding.

**Conclusions:** Shorter DAPT reduced major bleeding, and the ischemic cost of aspirin withdrawal declined with duration; a step at 1 month was not supported. A 15% excess of MI could be excluded from about 5 months (Emax) or 9 months (spline). The curves match duration to each patient’s ischemic margin.

**Registration:** URL: https://www.crd.york.ac.uk/PROSPERO/; Unique identifier: CRD420261480692.

**What is Known / What the Study Adds:** *What is Known:*

- Randomized trials have withdrawn aspirin after PCI at protocol-defined times from the index procedure to 6 months while continuing a P2Y12 inhibitor, and guidelines support this transition after 1 to 3 months in selected patients.
- Meta-analyses have compared these trials as duration categories and disagree on how short is safe.

*What the Study Adds:*

- With duration modeled as a continuous exposure, the ischemic cost of aspirin withdrawal declined with the preceding duration of DAPT and a step at 1 month was not supported, while major bleeding decreased continuously with shorter duration.
- Under every model the point estimate for myocardial infarction exceeded a 15% bound below about 3 weeks and fell below it from about 90 days; the credible bound was met at 5 to 9 months depending on the fitted function.
- How short is safe depends on the ischemic margin accepted for each patient, which these curves make explicit.

## INTRODUCTION

Dual antiplatelet therapy (DAPT) with aspirin and a P2Y12 inhibitor remains the default regimen after percutaneous coronary intervention (PCI), but the way DAPT ends has changed. In the contemporary paradigm it is aspirin that is withdrawn while the P2Y12 inhibitor continues as monotherapy, so that the duration of DAPT is, operationally, the duration of aspirin co-therapy. Randomized trials have tested this transition at protocol-defined time points ranging from the index procedure to several months, most often against 12 months of DAPT, and guidelines and consensus documents now support P2Y12 inhibitor monotherapy after 1 to 3 months in selected patients.^1–4^ The practical question facing clinicians has become not whether but *when* aspirin can be stopped.

Several syntheses published in 2026 addressed that question from a largely shared set of trials and reached different conclusions, favoring 1 month, 3 months, or no particular duration^5–8^; others compared ultrashort with standard-duration DAPT as two categories.^9^ These analyses differ mainly in how the exposure was categorized. A category labeled "≤1 month" pools trials that stopped aspirin within days of PCI with trials that stopped it at 30 days, and a comparison between two or three categories cannot show whether the ischemic cost of withdrawal changes as a step or as a slope. Whether a threshold exists, and where the cost becomes acceptably small, are questions about the shape of a continuous relation that categorical comparisons cannot answer.^10^

We therefore modeled the duration of DAPT before aspirin withdrawal as a continuous exposure in a dose-response network meta-analysis of randomized trials, with a prespecified estimand — the shortest duration at which an excess of myocardial infarction relative to 12-month DAPT can be excluded — and a parallel analysis of major bleeding to quantify the benefit retained at that duration.

## METHODS

### Registration and reporting

The protocol was registered in PROSPERO (CRD420261480692) before formal searching; its successive versions are listed in Supplementary Table S4. Reporting follows PRISMA 2020 and its extension for network meta-analysis.^11,12^ The study used aggregate data from published trials, so institutional review board approval and informed consent were not required.

### Eligibility criteria

We included randomized trials of adults undergoing PCI with drug-eluting stents for acute or chronic coronary syndromes in which the randomized comparison was the duration of DAPT, DAPT was terminated by withdrawing aspirin while a P2Y12 inhibitor (clopidogrel, prasugrel, or ticagrelor) was continued as monotherapy, and arm-level event counts were available at 12 months. Trials that terminated DAPT by withdrawing the P2Y12 inhibitor and continuing aspirin (ISAR-SAFE,^13^ ITALIC,^14^ and SECURITY^15^) were excluded. Trials that randomized patients only after 9 to 12 months or more of DAPT (OPT-BIRISK^16^; A-CLOSE^17^; DAPT-MVD^18^) lie outside the exposure axis and were excluded. MASTER DAPT^19^ was not used as a data point because the duration of DAPT in its comparator arm was not fixed and the agent continued after abbreviated therapy was left to physician discretion, and STOPDAPT-3^20^ was excluded because its comparator arm received 1 month of DAPT followed by aspirin monotherapy. HOST-BR,^21^ which randomized separate strata to different pairs of durations (1 versus 3 months in patients at high bleeding risk; 3 versus 12 months in others), contributed one analysis unit per stratum. Where a parent trial and a landmark substudy overlapped (GLOBAL LEADERS^22^ and GLASSY^23^), the parent was used.

### Exposure

The exposure was the protocol-assigned duration of DAPT before aspirin withdrawal, coded in days as 0 (aspirin withheld from the index procedure), 16, 30, 45, 90, 180, or 365 according to each trial’s design (Supplementary Table S2). Assigned duration was used in the base case because it is defined for every trial. Achieved duration, the median time from the index procedure to actual aspirin discontinuation, was extracted where reported or structurally determined, graded by a prespecified evidence hierarchy (A to D; Supplementary Methods), and used in the principal sensitivity analysis. For PREMIUM,^24^ whose control arm showed 56.5% aspirin persistence at 12 months, achieved duration was approximated from visit-wise persistence and entered as a range.

### Outcomes

The co-primary outcomes were myocardial infarction and Bleeding Academic Research Consortium (BARC) type 3 or 5 bleeding at 12 months, analyzed separately because ischemic and bleeding effects move in opposite directions along the duration axis. Where the BARC 3 or 5 composite was not published it was requested from the authors; where it could not be obtained (NEO-MINDSET^25^), BARC type 3 alone was used as a conservative substitute. Trials with a delayed observation window (SHARE,^26^ 3 to 12 months) or per-protocol reporting of ischemic outcomes (TWILIGHT^27^) were entered with their own denominators, which the study-specific baselines absorb. Secondary outcomes were definite or probable stent thrombosis, all-cause death, stroke, and each trial’s primary composite endpoint, reported as dose-response curves without a noninferiority bound because events are sparse or definitions differ (Supplementary Methods).

### Search and study selection

We searched PubMed, Embase, CENTRAL, ClinicalTrials.gov, and the WHO International Clinical Trials Registry Platform on August 18–20, 2026, without language or date restrictions, and checked the reference lists of recent reviews and meta-analyses^5–8,10^ (Supplementary Appendix S1). A top-up search of all sources on October 7, 2026 identified no additional eligible trial (Supplementary Appendix S1). After deduplication, records were filtered by prespecified keyword rules whose recall against 13 index trials was 13 of 13, and were then screened at title and abstract in two stages: a first pass by a large-language-model classifier applying the eligibility criteria, followed by independent dual screening of the candidate records by one reviewer (J.-S.J.) and a blinded second run of the classifier (κ = 0.78), with disagreements adjudicated by a second reviewer (K.-H.K.) and the adjudication verified by a third (J.S.P.). The false-negative rate of the automated steps was estimated by blinded dual re-screening of a random sample of 1,000 excluded records (Supplementary Methods). Full texts were assessed by two reviewers, and screening decisions were recorded with timestamps before any outcome data were extracted.

### Data extraction

Arm-level denominators, event counts, and assigned and achieved durations were extracted independently by two reviewers (J.-S.J. and J.S.P., the second blinded to the first) from the main report, supplementary appendix, and protocol of each trial, with the source of every value recorded; disagreements were resolved against the trial protocols. Where published reports did not permit non-overlapping inclusion or did not report an outcome in the required form, we contacted the trial authors; queries, replies, and their use are listed in Supplementary Table S6. Author-supplied stratum counts allowed STOPDAPT-2^28^ and STOPDAPT-2 ACS,^29^ whose populations overlap, to be included without double counting (Supplementary Methods).

### Risk of bias and certainty of evidence

Risk of bias for each co-primary outcome was assessed with RoB 2^30^ (effect of assignment) by two reviewers independently, with differences resolved against protocols, statistical analysis plans, and registry entries; events adjudicated by an independent blinded committee were not regarded as a measurement concern in open-label trials. Certainty of evidence was rated with the Grading of Recommendations Assessment, Development and Evaluation (GRADE) approach for network meta-analysis^31,32^ at prespecified positions on the exposure axis (0, 16, 30, 90, 158, and 180 days) and for the location of the estimand, judging imprecision against the prespecified threshold^33^ and indirectness with explicit attention to landmark (event-free) randomization, discretionary monotherapy agents, and residual uncertainty in achieved duration^34^; the ratings and a summary of findings are given in Supplementary Table S8. Risk of bias due to missing results was judged within the GRADE publication-bias domain from the registry searches and the registration status of the included trials; funnel-plot and regression-based tests were not used because the comparisons lie at different positions on the exposure axis rather than repeating one contrast.

### Statistical analysis

We used Bayesian model-based network meta-analysis (MBNMA), which treats each randomized arm as an observation at a known dose and fits a dose-response function across trials while preserving the randomized comparison within each trial.^35,36^ Models were fitted with the MBNMAdose package (version 0.5.1) in R and JAGS,^37^ with a binomial likelihood and logit link for arm-level event counts, DAPT duration in days as the dose of a single agent, study-specific baselines, and random treatment effects with weakly informative priors (Supplementary Methods). The model assumes one dose-response relation shared by all trials, with the random effect absorbing between-trial differences in the size of the effect; differences in clinical presentation, monotherapy agent, and landmark versus index-procedure randomization were therefore examined as potential effect modifiers in the sensitivity analyses rather than built into the primary curve. Two smooth functions were prespecified, an Emax function and a restricted cubic spline with knots at 16 and 90 days, with change-point models at 30 or 90 days as prespecified alternatives. Each model was run for 60,000 iterations in three chains after burn-in, with convergence required for every parameter (potential scale reduction factor <1.05); fit was compared by the deviance information criterion (DIC) and residual deviance, and inconsistency between the dose-response assumption and the data was checked against an unrelated-mean-effects model. Odds ratios (ORs) versus 12-month DAPT were computed from the posterior samples as differences of fitted logits.

The prespecified estimand was the shortest duration at which the posterior upper 95% credible limit of the OR for myocardial infarction versus 12-month DAPT is below 1.15, an OR-scale application of the hazard-ratio bound registered in the protocol. Because the credible bound proved non-monotone under the spline, we report both the first duration at which the bound is met and the earliest duration from which it is met continuously to 365 days, and use the latter as the principal value; and when the two functions differed in DIC by less than 3, within the Monte Carlo noise observed between replicate runs, the more parsimonious Emax function was used for the primary curve; both rules were adopted after the first model fits (Supplementary Table S4). Bleeding curves are reported without a bound. Because the Monte Carlo error of the estimand was comparable to the 7-day agreement rule described below, all estimands were computed from the 60,000-iteration runs.

Sensitivity analyses are numbered as in the protocol, in which eight (S1 to S8) were prespecified; Supplementary Table S7 lists them all. The principal sensitivity analysis, S6, designated as such before analysis (Supplementary Table S4), coded units at achieved rather than assigned duration, with agreement defined a priori as a difference of 7 days or less in the estimand, and was run in three forms (Supplementary Methods). S4 fitted the change-point models, and S5 recomputed the estimand under margins equivalent to absolute risk differences of 0.5, 1.0, and 1.5 percentage points at the pooled control-arm risk. Network meta-regressions were specified for region (S3), calendar period (S7), monotherapy agent (S1), landmark design (S2), and protocol-fixed versus discretionary agent (S8); because the regression term is identified only when both levels of a covariate include an arm at 0 days, S1 and S2 were replaced by stratified and exclusion analyses and S8 could not be estimated (Supplementary Table S7). Leave-one-out analyses were run for each trial contributing two units or a single exposure level and for the trial with the largest share of events, and an alternative partition of the STOPDAPT-2 trials by clinical presentation was examined. Two further analyses were not prespecified (Supplementary Table S4): S9 used author-supplied data to restrict the HOST-BR monotherapy arms to patients prescribed a P2Y12 inhibitor, and S10 examined clinical presentation by a meta-regression on the proportion of patients with acute coronary syndrome and by stratifying trials that enrolled acute coronary syndrome only. All analyses used a fixed seed.

### Data sharing

The extracted data, analysis code, and model outputs are deposited at Zenodo (https://doi.org/10.5281/zenodo.23250940) and will be made publicly available at publication. J.-S.J. had full access to all the data in the study and takes responsibility for its integrity and the data analysis.

## RESULTS

### Study selection and characteristics

The searches identified 21,725 records; after deduplication, keyword filtering, and screening of 2,898 records, 293 reports were assessed in full text and 17 trials met the eligibility criteria (Supplementary Figure S1). Sixteen were synthesized,^21,22,24–29,38–45^ because the remaining trial reported bleeding in a form from which first-event counts could not be recovered (Supplementary Table S3). No eligible study was missed in the blinded re-screen of 1,000 records excluded by the automated steps.

The 16 trials, published between 2018 and 2026, contributed 17 analysis units and 58,068 patients, with 929 myocardial infarctions and 1,102 BARC 3 or 5 bleeding events at 12 months (Table 1); the myocardial infarction network comprised 57,988 patients because TWILIGHT reported ischemic outcomes in its per-protocol population. Ten units were conducted in East Asia, nine enrolled patients with acute coronary syndrome exclusively, and three randomized patients after an event-free interval (landmark design). The agent continued as monotherapy was ticagrelor in six units, prasugrel in three, clopidogrel in three, and left to the physician in five; in HOST-BR, 22% of patients assigned to monotherapy received aspirin rather than a P2Y12 inhibitor. The assigned duration of DAPT before aspirin withdrawal was 0 days in three units, 16 days in one, 30 days in seven, 45 days in one, 90 days in five, and 180 days in one, and the comparator was 12-month DAPT in all but the HOST-BR high-bleeding-risk stratum (3 months). On the exposure axis the units cluster at 0, 30, and 90 days, with single units at 16, 45, and 180 days; the 180-day unit is the only one between 90 and 365 days (Supplementary Figure S2).

**Table 1.**
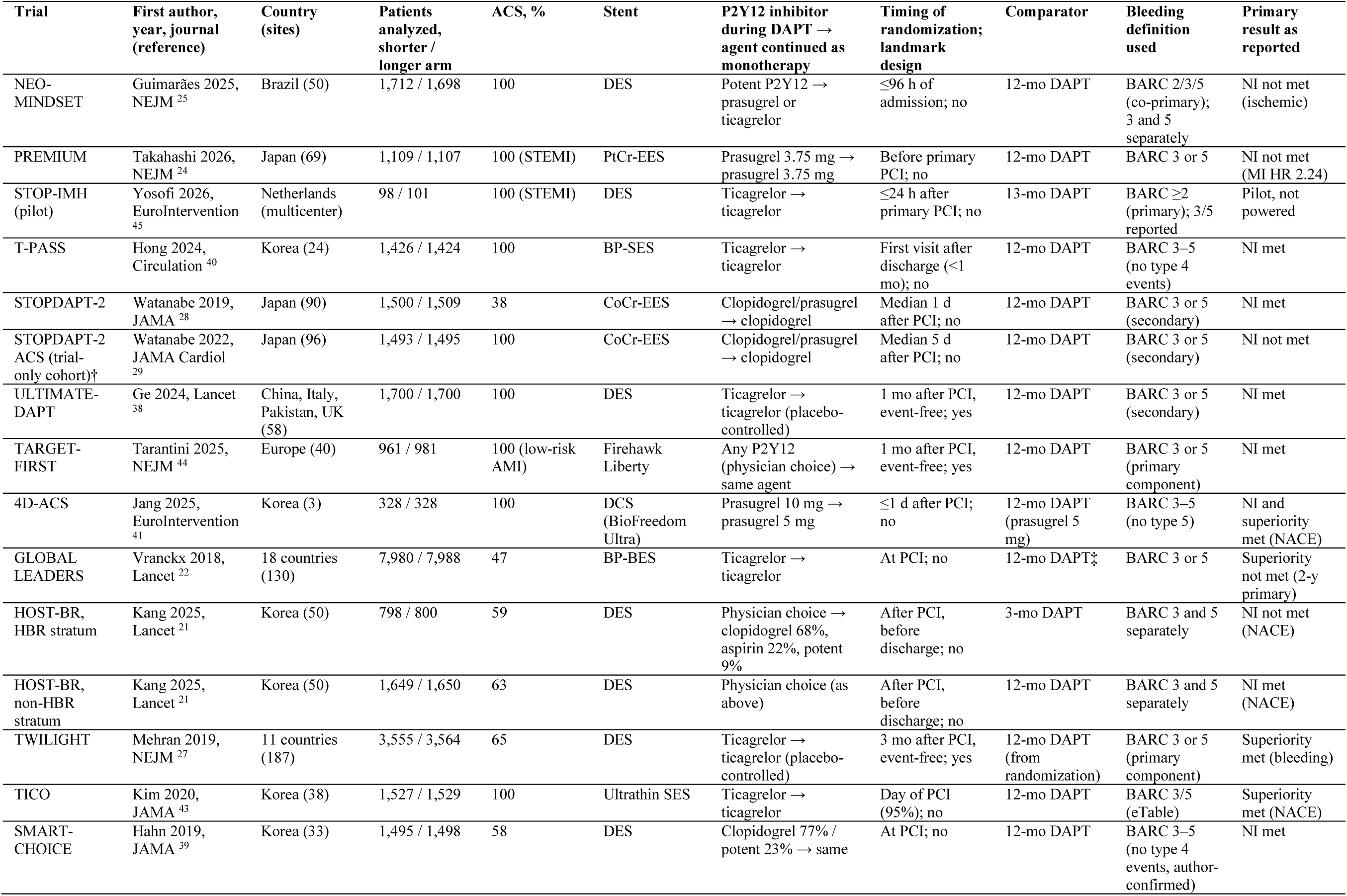
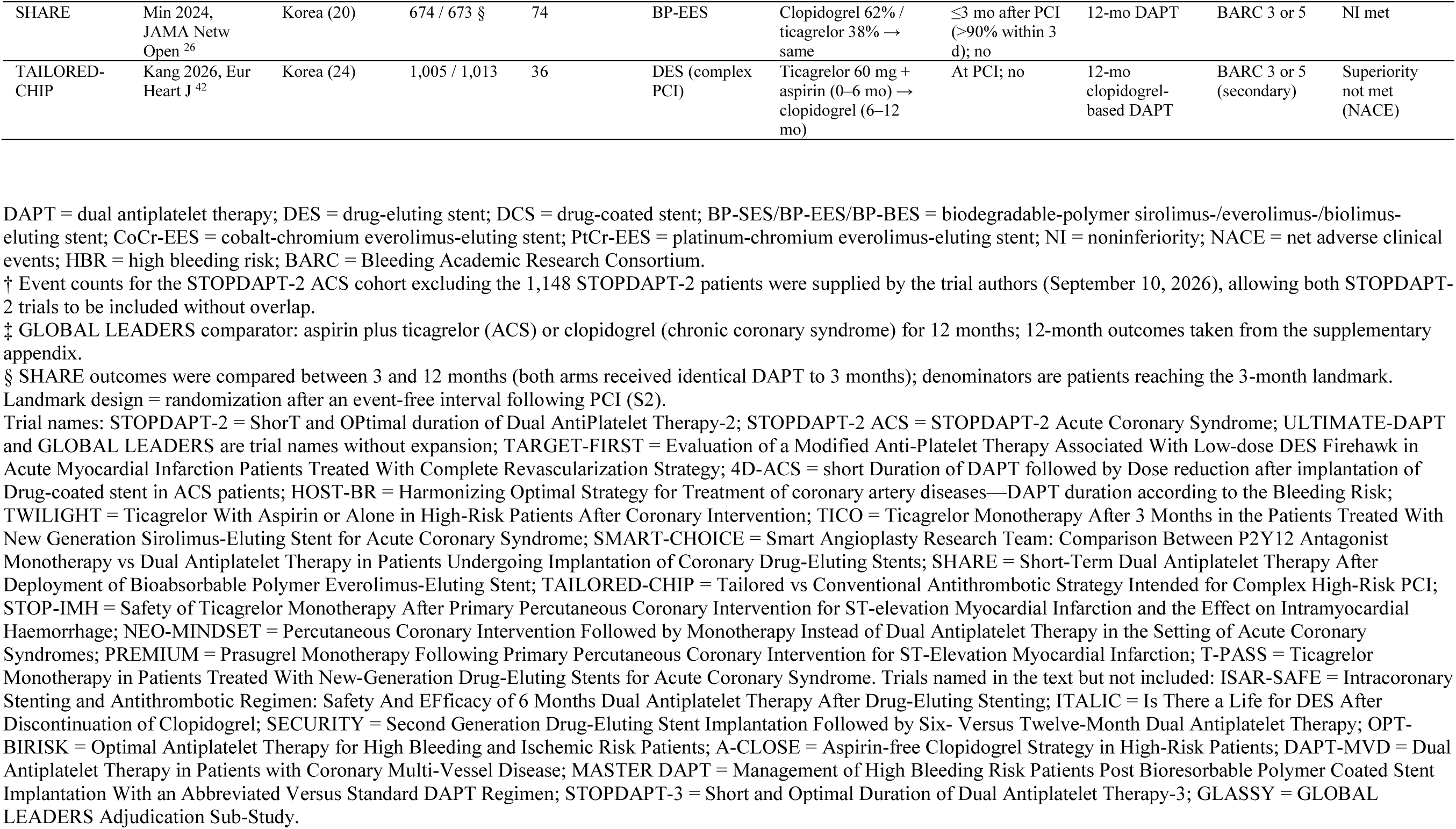
Characteristics of the Included Trials (16 Trials, 17 Analysis Units)

| Trial | First author, year, journal (reference) | Country (sites) | Patients analyzed, shorter / longer arm | ACS, % | Stent | P2Y12 inhibitor during DAPT → agent continued as monotherapy | Timing of randomization; landmark design | Comparator | Bleeding definition used | Primary result as reported |
| --- | --- | --- | --- | --- | --- | --- | --- | --- | --- | --- |
| NEO-MINDSET | Guimarães 2025, NEJM <sup>25</sup> | Brazil (50) | 1,712 / 1,698 | 100 | DES | Potent P2Y12 → prasugrel or ticagrelor | ≤96 h of admission; no | 12-mo DAPT | BARC 2/3/5 (co-primary); 3 and 5 separately | NI not met (ischemic) |
| PREMIUM | Takahashi 2026, NEJM <sup>24</sup> | Japan (69) | 1,109 / 1,107 | 100 (STEMI) | PtCr-EES | Prasugrel 3.75 mg → prasugrel 3.75 mg | Before primary PCI; no | 12-mo DAPT | BARC 3 or 5 | NI not met (MI HR 2.24) |
| STOP-IMH (pilot) | Yosofi 2026, EuroIntervention <sup>45</sup> | Netherlands (multicenter) | 98 / 101 | 100 (STEMI) | DES | Ticagrelor → ticagrelor | ≤24 h after primary PCI; no | 13-mo DAPT | BARC ≥2 (primary); 3/5 reported | Pilot, not powered |
| T-PASS | Hong 2024, Circulation <sup>40</sup> | Korea (24) | 1,426 / 1,424 | 100 | BP-SES | Ticagrelor → ticagrelor | First visit after discharge (<1 mo); no | 12-mo DAPT | BARC 3–5 (no type 4 events) | NI met |
| STOPDAPT-2 | Watanabe 2019, JAMA <sup>28</sup> | Japan (90) | 1,500 / 1,509 | 38 | CoCr-EES | Clopidogrel/prasugrel → clopidogrel | Median 1 d after PCI; no | 12-mo DAPT | BARC 3 or 5 (secondary) | NI met |
| STOPDAPT-2 ACS (trial-only cohort)† | Watanabe 2022, JAMA Cardiol <sup>29</sup> | Japan (96) | 1,493 / 1,495 | 100 | CoCr-EES | Clopidogrel/prasugrel → clopidogrel | Median 5 d after PCI; no | 12-mo DAPT | BARC 3 or 5 (secondary) | NI not met |
| ULTIMATE-DAPT | Ge 2024, Lancet <sup>38</sup> | China, Italy, Pakistan, UK (58) | 1,700 / 1,700 | 100 | DES | Ticagrelor → ticagrelor (placebo-controlled) | 1 mo after PCI, event-free; yes | 12-mo DAPT | BARC 3 or 5 (secondary) | NI met |
| TARGET-FIRST | Tarantini 2025, NEJM <sup>44</sup> | Europe (40) | 961 / 981 | 100 (low-risk AMI) | Firehawk Liberty | Any P2Y12 (physician choice) → same agent | 1 mo after PCI, event-free; yes | 12-mo DAPT | BARC 3 or 5 (primary component) | NI met |
| 4D-ACS | Jang 2025, EuroIntervention <sup>41</sup> | Korea (3) | 328 / 328 | 100 | DCS (BioFreedom Ultra) | Prasugrel 10 mg → prasugrel 5 mg | ≤1 d after PCI; no | 12-mo DAPT (prasugrel 5 mg) | BARC 3–5 (no type 5) | NI and superiority met (NACE) |
| GLOBAL LEADERS | Vranckx 2018, Lancet <sup>22</sup> | 18 countries (130) | 7,980 / 7,988 | 47 | BP-BES | Ticagrelor → ticagrelor | At PCI; no | 12-mo DAPT‡ | BARC 3 or 5 | Superiority not met (2-y primary) |
| HOST-BR, HBR stratum | Kang 2025, Lancet <sup>21</sup> | Korea (50) | 798 / 800 | 59 | DES | Physician choice → clopidogrel 68%, aspirin 22%, potent 9% | After PCI, before discharge; no | 3-mo DAPT | BARC 3 and 5 separately | NI not met (NACE) |
| HOST-BR, non-HBR stratum | Kang 2025, Lancet <sup>21</sup> | Korea (50) | 1,649 / 1,650 | 63 | DES | Physician choice (as above) | After PCI, before discharge; no | 12-mo DAPT | BARC 3 and 5 separately | NI met (NACE) |
| TWILIGHT | Mehran 2019, NEJM <sup>27</sup> | 11 countries (187) | 3,555 / 3,564 | 65 | DES | Ticagrelor → ticagrelor (placebo-controlled) | 3 mo after PCI, event-free; yes | 12-mo DAPT (from randomization) | BARC 3 or 5 (primary component) | Superiority met (bleeding) |
| TICO | Kim 2020, JAMA <sup>43</sup> | Korea (38) | 1,527 / 1,529 | 100 | Ultrathin SES | Ticagrelor → ticagrelor | Day of PCI (95%); no | 12-mo DAPT | BARC 3/5 (eTable) | Superiority met (NACE) |
| SMART-CHOICE | Hahn 2019, JAMA <sup>39</sup> | Korea (33) | 1,495 / 1,498 | 58 | DES | Clopidogrel 77% / potent 23% → same | At PCI; no | 12-mo DAPT | BARC 3–5 (no type 4 events, author-confirmed) | NI met |
| SHARE | Min 2024, JAMA Netw Open <sup>26</sup> | Korea (20) | 674 / 673 § | 74 | BP-EES | Clopidogrel 62% / ticagrelor 38% → same | ≤3 mo after PCI (>90% within 3 d); no | 12-mo DAPT | BARC 3 or 5 | NI met |
| TAILORED-CHIP | Kang 2026, Eur Heart J <sup>42</sup> | Korea (24) | 1,005 / 1,013 | 36 | DES (complex PCI) | Ticagrelor 60 mg + aspirin (0–6 mo) → clopidogrel (6–12 mo) | At PCI; no | 12-mo clopidogrel-based DAPT | BARC 3 or 5 (secondary) | Superiority not met (NACE) |

DAPT = dual antiplatelet therapy; DES = drug-eluting stent; DCS = drug-coated stent; BP-SES/BP-EES/BP-BES = biodegradable-polymer sirolimus-/everolimus-/biolimus-eluting stent; CoCr-EES = cobalt-chromium everolimus-eluting stent; PtCr-EES = platinum-chromium everolimus-eluting stent; NI = noninferiority; NACE = net adverse clinical events; HBR = high bleeding risk; BARC = Bleeding Academic Research Consortium.
† Event counts for the STOPDAPT-2 ACS cohort excluding the 1,148 STOPDAPT-2 patients were supplied by the trial authors (September 10, 2026), allowing both STOPDAPT-2 trials to be included without overlap.
‡ GLOBAL LEADERS comparator: aspirin plus ticagrelor (ACS) or clopidogrel (chronic coronary syndrome) for 12 months; 12-month outcomes taken from the supplementary appendix.
§ SHARE outcomes were compared between 3 and 12 months (both arms received identical DAPT to 3 months); denominators are patients reaching the 3-month landmark.
Landmark design = randomization after an event-free interval following PCI (S2).

Author-supplied data resolved the overlap between the STOPDAPT-2 trials, the BARC 3 or 5 composite for HOST-BR, and the absence of BARC type 4 events in SMART-CHOICE^39^ (Supplementary Table S6). Achieved duration was available for 15 of 17 units (Supplementary Table S2).

### Risk of bias

Risk of bias was low in nine units for myocardial infarction and in eight for bleeding, with some concerns in the remainder, mostly for selective reporting where a dated protocol was unavailable or the bleeding definition was post hoc, and high in one unit (SHARE), in which 4.5% of randomized patients were excluded before the 3-month landmark (Supplementary Table S5).

### Dose-response for myocardial infarction

Both smooth functions fitted the data adequately (Emax: DIC 208.6, residual deviance 34.0 for 34 arms; spline: DIC 208.2, residual deviance 32.9; between-study SD 0.18 on the log-odds scale), and neither differed materially from the unrelated-mean-effects model (DIC 206.6 and 204.6), so the Emax function was taken as primary. The Emax curve decreased monotonically with duration (Figure 1, Table 2). Relative to 12-month DAPT, the OR for myocardial infarction was 1.31 (95% credible interval [CrI] 0.95–2.09) with aspirin withdrawn at PCI, 1.21 (0.95–1.53) at 16 days, 1.17 (0.96–1.43) at 30 days, 1.09 (0.98–1.25) at 90 days, and 1.04 (0.99–1.13) at 180 days. The median OR fell below 1.15 at 39 days, but the upper credible limit did so only at 158 days (OR 1.05, 95% CrI 0.99–1.15), from which it remained below the bound to 365 days; the two readings of the estimand coincided. The spline was steeper at short durations (OR 1.55, 0.98–2.40, at 0 days; 1.33, 1.01–1.74, at 16 days) and dipped below 1 between 60 and 300 days; its upper limit first met the bound at 82 days but re-crossed it between about 120 and 280 days, an interval informed by a single trial at 180 days, so that the continuously satisfied reading was 282 days. Because the two functions fitted equally, the credible bound is best read as a range from 158 to 282 days. Two findings did not depend on the function: under the Emax, spline, and both change-point models the median OR exceeded 1.15 at every duration below about 3 weeks and was 1.09 or lower from 90 days onward.

**Figure 1.**
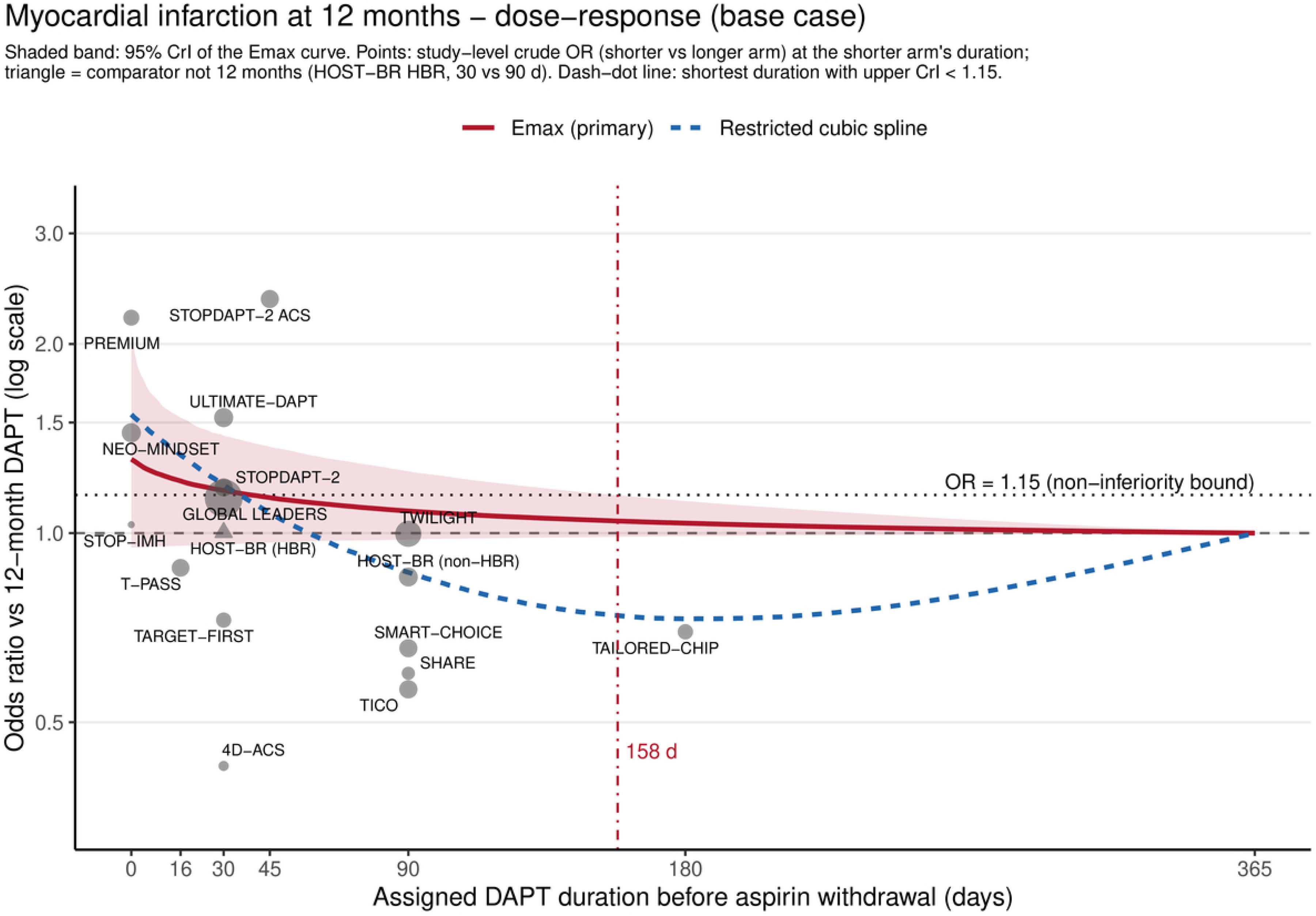
Dose-response relation between the assigned duration of DAPT before aspirin withdrawal and myocardial infarction at 12 months. Odds ratios versus 12-month DAPT (log scale) from the Emax model (red, primary; shaded band = 95% credible interval) and the restricted cubic spline (blue dashed) in 17 randomized comparisons (58,068 patients). Points are study-level crude odds ratios of the shorter versus the longer arm, plotted at the shorter arm’s assigned duration and sized by trial size; the triangle marks the HOST-BR high-bleeding-risk stratum, whose comparator was 3 months. The dotted horizontal line is the prespecified noninferiority bound (odds ratio 1.15); the dash-dot vertical line marks 158 days, the shortest duration at which the upper credible limit of the Emax curve remains below the bound to 365 days.

**Table 2.** Odds Ratios Versus 12-Month DAPT Along the Duration Axis and the Prespecified Estimand.

| **Duration of DAPT before aspirin withdrawal** | **Myocardial infarction, Emax (primary)** | **Myocardial infarction, spline** | **BARC 3 or 5 bleeding, Emax (primary)** | **BARC 3 or 5 bleeding, spline** |
| --- | --- | --- | --- | --- |
| 0 days (at PCI) | 1.31 (0.95–2.09) | 1.55 (0.98–2.40) | 0.40 (0.26–0.56) | 0.46 (0.23–0.84) |
| 7 days | 1.25 (0.95–1.66) | 1.44 (1.02–2.01) | 0.42 (0.28–0.58) | 0.46 (0.27–0.70) |
| 16 days | 1.21 (0.95–1.53) | 1.33 (1.01–1.74) | 0.45 (0.31–0.60) | 0.45 (0.30–0.62) |
| 30 days | 1.17 (0.96–1.43) | 1.19 (0.87–1.59) | 0.48 (0.34–0.63) | 0.45 (0.29–0.65) |
| 45 days | 1.14 (0.97–1.37) | 1.08 (0.76–1.45) | 0.52 (0.38–0.67) | 0.46 (0.29–0.70) |
| 90 days | 1.09 (0.98–1.25) | 0.87 (0.64–1.12) | 0.62 (0.49–0.76) | 0.54 (0.35–0.79) |
| 158 days (estimand) | 1.05 (0.99–1.15) | 0.74 (0.44–1.19) | 0.74 (0.63–0.86) | 0.67 (0.35–1.32) |
| 180 days | 1.04 (0.99–1.13) | 0.73 (0.43–1.21) | 0.77 (0.68–0.88) | 0.72 (0.36–1.46) |
| 270 days | 1.01 (1.00–1.05) | 0.80 (0.54–1.17) | 0.90 (0.84–0.95) | 0.87 (0.52–1.49) |
| **Estimand: shortest duration with upper 95% CrI of the MI OR < 1.15** |  |  |  |  |
| — met continuously to 365 days (principal) | 158 days | 282 days | — | — |
| — first met | 158 days | 82 days | — | — |
| — achieved-duration coding (S6b / S6a / S6c) | 166 / 174 / 158 days | 84 / 94 / 83 days | — | — |
| Model fit: DIC (residual deviance / arms) | 208.6 (34.0/34) | 208.2 (32.9/34) | 224.9 (34.1/34) | 225.7 (34.0/34) |
| Unrelated-mean-effects model, DIC | 206.6 | 204.6 | — | — |

Values are posterior median odds ratios (95% credible intervals) versus 12-month DAPT (365 days), from Bayesian model-based network meta-analysis of 17 analysis units (16 trials; 58,068 patients; 929 myocardial infarctions; 1,102 BARC 3 or 5 bleeding events). The spline is a natural cubic spline with knots at 16 and 90 days. Abbreviations: BARC = Bleeding Academic Research Consortium; CrI = credible interval; DAPT = dual antiplatelet therapy; DIC = deviance information criterion; MI = myocardial infarction; PCI = percutaneous coronary intervention.

### Dose-response for major bleeding

For BARC 3 or 5 bleeding the Emax function was again primary (DIC 224.9 versus 225.7; between-study SD 0.39; unrelated-mean-effects DIC 231.7). Bleeding decreased continuously with shorter duration without a plateau (Figure 2, Table 2): OR 0.40 (95% CrI 0.26–0.56) at 0 days, 0.48 (0.34–0.63) at 30 days, 0.62 (0.49–0.76) at 90 days, and 0.77 (0.68–0.88) at 180 days, with the upper credible limit below 1 at every duration to 364 days. At 158 days the OR for bleeding was 0.74 (0.63–0.86). The spline gave similar values at short durations, with wider intervals over the sparsely sampled middle of the axis.

**Figure 2.**
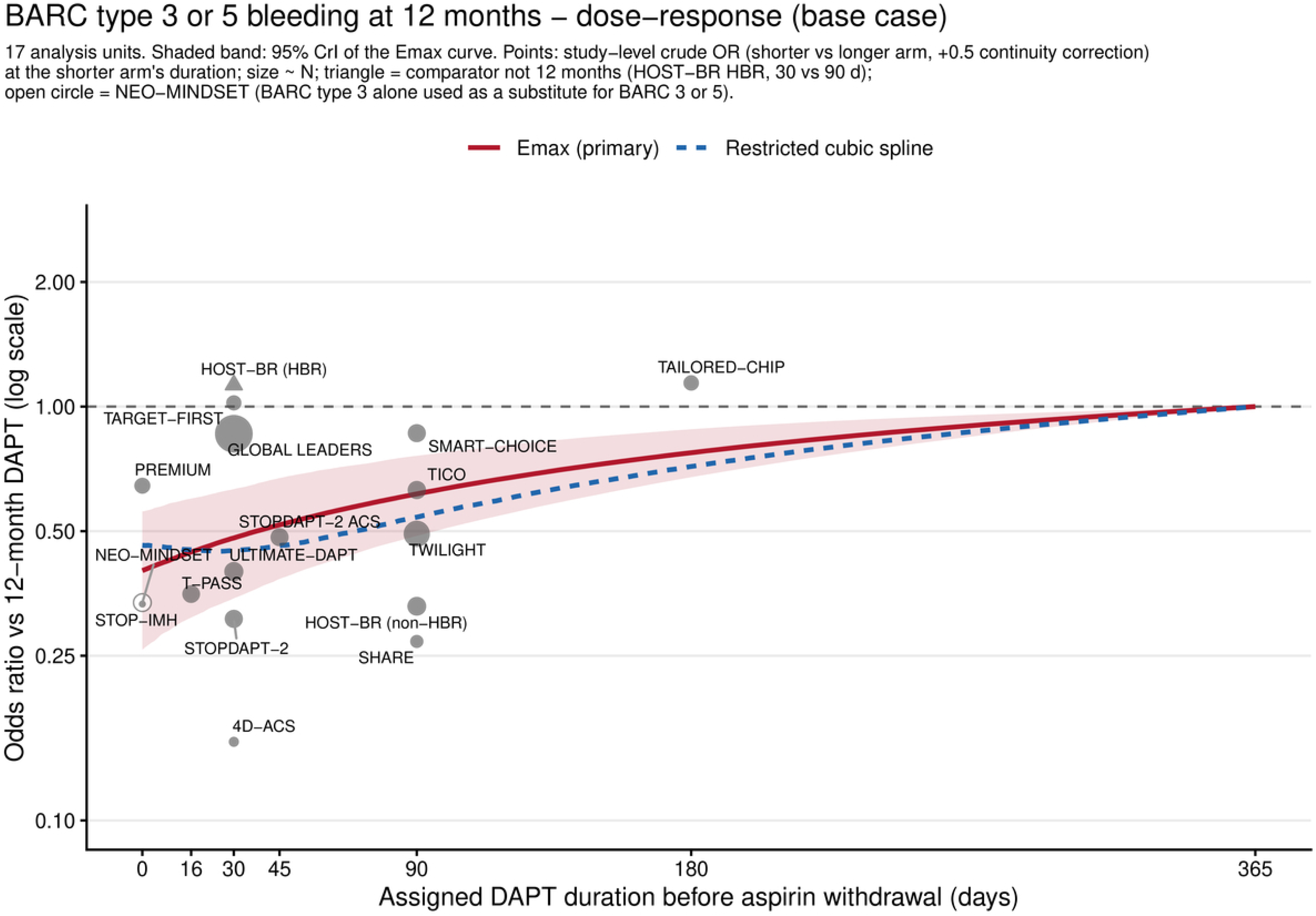
Dose-response relation between the assigned duration of DAPT before aspirin withdrawal and BARC type 3 or 5 bleeding at 12 months. Odds ratios versus 12-month DAPT (log scale) from the Emax model (red, primary; shaded band = 95% credible interval) and the restricted cubic spline (blue dashed) in 17 randomized comparisons, including both HOST-BR strata with author-supplied composite counts. Points are study-level crude odds ratios of the shorter versus the longer arm (0.5 added to each cell), plotted at the shorter arm’s assigned duration and sized by trial size; the triangle marks the HOST-BR high-bleeding-risk stratum, whose comparator was 3 months, and the open circle marks NEO-MINDSET, for which BARC type 3 alone was used as a substitute for BARC 3 or 5. No noninferiority bound applies to bleeding.

### Achieved versus assigned duration

Coding units at achieved rather than assigned duration moved the Emax estimand from 158 to 166 days when the two units without usable data retained their assigned values (S6b), to 174 days when they were excluded (S6a), and left it at 158 days with PREMIUM at the assigned end of its range (S6c; Supplementary Figure S3). The 8-day difference between the base case and S6b lies at the prespecified 7-day boundary and is of the same order as the Monte Carlo error between replicate runs (about 5 days); we therefore report both values as approximately 5 months. Under the spline, achieved-duration coding shortened the first crossing to 84 days without altering the non-monotone bound.

### Other sensitivity analyses

The change-point models did not fit better than the smooth functions (DIC 210.2 with a knot at 30 days, 207.2 at 90 days; Supplementary Figure S4). The estimand was highly sensitive to the noninferiority margin (Supplementary Figure S5): margins equivalent to absolute risk differences of 0.5, 1.0, and 1.5 percentage points at the pooled control-arm risk of 1.5% (ORs 1.33, 1.67, and 2.01) gave 58, 7, and 1 days. Meta-regressions for region (log-OR interaction −0.68, 95% CrI −1.58 to 0.20) and calendar period (−0.12 per year, −0.35 to 0.10) did not indicate effect modification; those for monotherapy agent, landmark design, and protocol-fixed agent could not be identified, and the corresponding stratified analyses were underpowered (Supplementary Table S7). Excluding the three landmark-design units lengthened the estimand to 187 days.

Removing single trials moved the estimand between 133 and 179 days (Figure 3): to 133 days without STOPDAPT-2 ACS, whose 45-day arm had an OR of 2.4, and to 179 days without GLOBAL LEADERS, whose 30-day arm (OR 1.14, 337 events) anchors that node near the null; removal of HOST-BR, TAILORED-CHIP,^42^ STOPDAPT-2, or T-PASS^40^ gave 161 to 172 days, and the alternative partition of the STOPDAPT-2 trials gave 158 days. Restricting the HOST-BR monotherapy arms to patients prescribed a P2Y12 inhibitor (S9) gave 160 days. In the nine units that enrolled acute coronary syndrome only, which include every unit below 30 days, the curve was steeper at every position (OR 1.29, 95% CrI 0.87–1.90, at 30 days; 1.16, 0.88–1.53, at 90 days) and the estimand was 238 days (S10); the eight mixed-presentation units contain no arm below 30 days, and no trial enrolled chronic coronary syndrome only, so a presentation-specific comparison could not be made and the meta-regression on the proportion with acute coronary syndrome did not converge (Supplementary Table S7).

**Figure 3.**
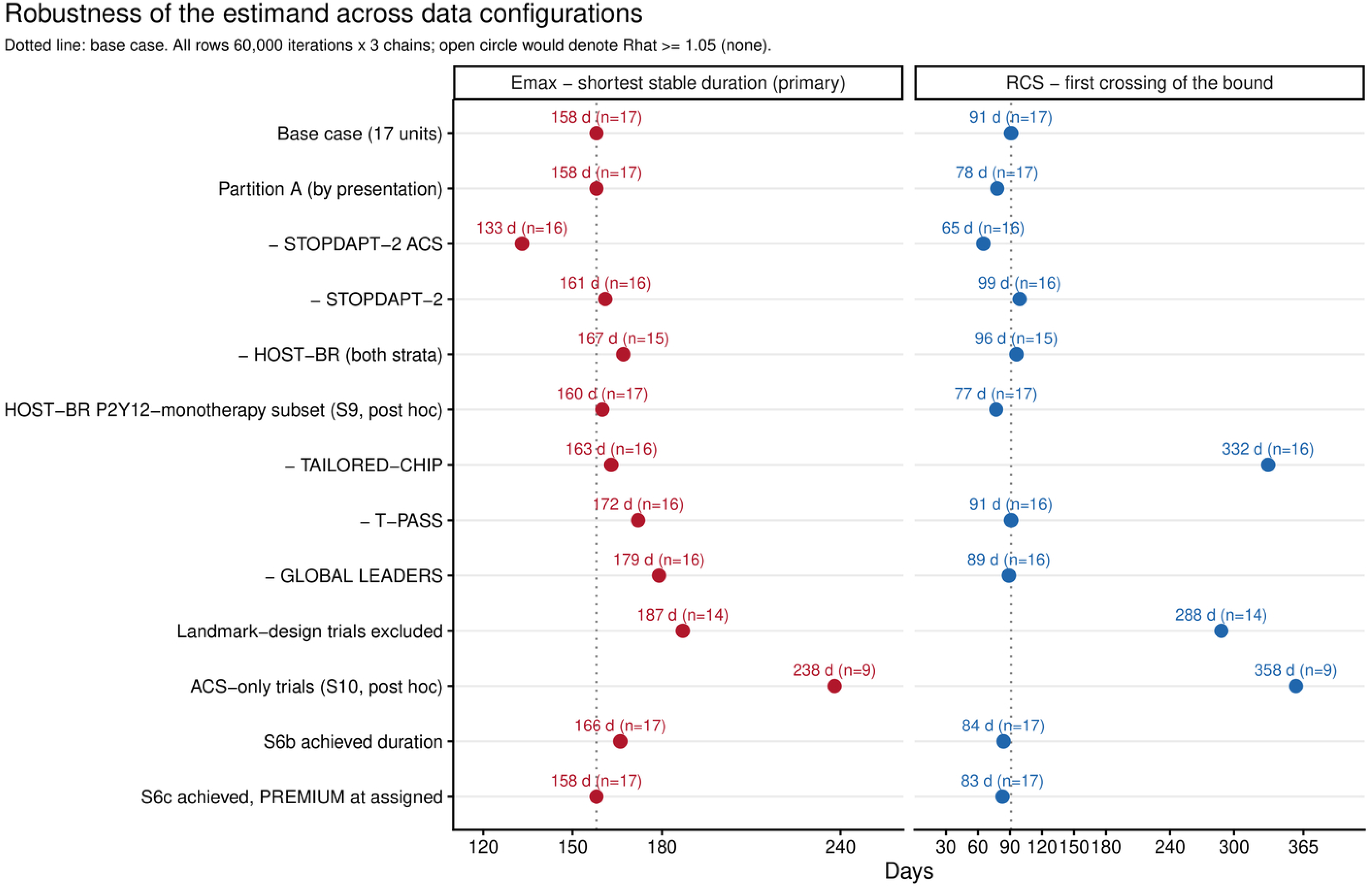
Robustness of the estimand across data configurations. Shortest duration at which the upper 95% credible limit of the odds ratio for myocardial infarction is below 1.15 and remains so to 365 days (Emax, left) and first crossing of the bound (spline, right) in the base case, the alternative partition of the STOPDAPT-2 trials, leave-one-out analyses, the post hoc HOST-BR P2Y12-monotherapy subset (S9), exclusion of landmark-design units, the post hoc stratum of trials enrolling acute coronary syndrome only (S10), and the achieved-duration analyses (S6b, S6c). Dotted line = base case. Leaving out STOPDAPT-2 ACS removes the unit with the largest early excess of myocardial infarction and shortens the estimand; the acute-coronary-syndrome stratum retains that unit and omits the 8 mixed-presentation units, so its longer estimand reflects fewer events and a wider credible bound as much as a steeper curve. All models 60,000 iterations × 3 chains; no model had a potential scale reduction factor ≥1.05.

### Secondary outcomes

Curves were flat for all-cause death (OR 1.04, 95% CrI 0.82–1.36, at 0 days; 949 deaths) and stroke (1.03, 0.64–1.63; 442 events). Definite or probable stent thrombosis followed the direction of myocardial infarction with wide uncertainty (1.39, 0.80–2.87, at 0 days; 1.15, 0.90–1.64, at 90 days; 208 events), and each trial’s primary composite endpoint favored shorter duration with the upper credible limit at the null (0.75, 0.53–1.01, at 0 days; 0.86, 0.71–1.01, at 90 days). The spline did not converge for stroke or stent thrombosis (Supplementary Figure S6).

### Certainty of evidence

Certainty in the myocardial infarction curve was very low at 0 days (three trials, 79 events, credible interval spanning no effect to a doubling of risk), low at 16 to 90 days (credible intervals crossing the threshold; landmark designs and discretionary agents), and moderate at 158 to 180 days, downgraded once because no trial contributes between 90 and 180 days. Certainty in the location of the estimand was low, because it depends on the form of the function across the sparsely sampled interval between 90 and 365 days and moves by up to 25 days with single trials. Certainty in the bleeding curve was moderate at every position, downgraded once for indirectness; precision was adequate throughout, and neither heterogeneity, risk of bias, publication bias, nor incoherence was rated down (Supplementary Table S8).

## DISCUSSION

In 16 randomized trials that ended DAPT by withdrawing aspirin and continuing a P2Y12 inhibitor, the odds of myocardial infarction relative to 12-month DAPT declined as aspirin co-therapy lengthened (1.31 at the index procedure, 1.09 at 3 months), and a step at 1 month was not supported. Two findings did not depend on the function fitted: below about 3 weeks every model placed the point estimate above the prespecified 15% bound, and from about 90 days every model placed it below. Where the upper credible limit meets the bound did depend on the function: at 158 days with the Emax curve (166 by achieved duration) and at 282 days with the equally well-fitting spline, so that the estimand is a range of about 5 to 9 months rather than a date. Major bleeding fell continuously with shorter duration, by 60% at the index procedure, 38% at 3 months, and 26% at 5 months. Certainty was low for the ischemic curve between 16 and 90 days and for the location of the estimand (Supplementary Table S8). The finding is not that 5 months or more of DAPT are required, but that present evidence cannot exclude a 15% excess of myocardial infarction with less, and that "how short is safe" depends on a margin that categorical syntheses left implicit.

Why, then, has the field concluded that 1 month is enough? Our estimand asks when a 15% excess can be excluded, not when the point estimate falls below it. Under the margins the trials used, absolute differences of 1 to 2.7 percentage points or a relative margin of 50%, the same curve is satisfied within a week (Supplementary Figure S5). The 2026 meta-analyses drew on the same trials: a categorical comparison inherits whatever margin the reader brings to it, and the trials had normalized wide margins.^5–7^ "One month is enough" and "5 months are needed" are the same result read at different thresholds. The trials’ margins were chosen for feasibility, but the conclusion drawn from them is a statement about the evidence accepted rather than the biology observed. A recent state-of-the-art review made the point qualitatively^10^; the curve makes it quantitative.

The function-independent findings rest on the data rather than the model: the point estimates above the bound before 3 weeks come from the trials that withdrew aspirin at or within days of PCI, and those below it from 90 days come from the five units at 3 months. A step at 30 days, the implicit model behind the "1-month DAPT" label, was not supported: a change-point model with a knee at 30 days fitted no better than the Emax function, and a knee at 90 days fitted better only within the Monte Carlo noise of the deviance information criterion (Supplementary Figure S4).

Pharmacology predicts as much. Platelet cyclooxygenase-1 activity recovers over 5 to 7 days after aspirin is stopped,^46^ so withdrawal is itself graded, and the thrombotic hazard after stenting declines progressively rather than at a fixed date.^10^ The spline’s U-shaped curve, in which intermediate durations appeared safer than 12 months, is an artifact of the sparse region between 90 and 365 days, anchored by a single trial at 180 days (Supplementary Figure S2); it disappeared when the achieved-duration analysis placed a data point there (Supplementary Figure S3).

The trials are not interchangeable points on the axis, and the primary curve assumes that they share one dose-response relation. Three units randomized patients only after an event-free interval of 1 to 3 months. Their populations exclude early events, so withdrawal should look safer at 1 and 3 months than in all-comers; accordingly, removing the three landmark units lengthened the estimand to 187 days, the more conservative reading for a decision made at the index procedure. Single trials moved the estimand in both directions (Figure 3). Removing GLOBAL LEADERS, which supplies 36% of all myocardial infarctions and anchors the 1-month node near the null, lengthened it to 179 days. Removing STOPDAPT-2 ACS, the only trial at 45 days, with an odds ratio of 2.4, shortened it to 133 days; its influence reflects its position on the axis as much as its magnitude, and its own failure of noninferiority is consistent with the curve. The HOST-BR monotherapy arms, in which 22% of patients continued aspirin, did not pull the curve toward the null: restricting them to patients prescribed a P2Y12 inhibitor gave 160 days. Region and calendar period did not detectably modify the effect, although the interval for region (log-odds interaction −0.68, 95% CrI −1.58 to 0.20) does not exclude a smaller ischemic cost in East Asian populations, which contributed 10 of 17 units. Trials enrolling acute coronary syndrome only, which supply every unit below 30 days, gave a steeper curve and an estimand of 238 days; because those trials also used potent P2Y12 inhibitors and were mostly East Asian, and no trial enrolled chronic coronary syndrome only, the analysis indicates that the short-duration harm is driven by the acute-presentation trials without isolating presentation itself as the cause. Taken together, the analyses excluding landmark units (187 days) and restricted to acute coronary syndrome trials (238 days) show that the common curve is an average over populations and designs whose own curves differ, and the estimand should be read with that assumption in view.

The reference point of these estimates is conventional, not optimal: every estimate is relative to 12 months of DAPT because that is the comparator the trials share. A-CLOSE is a reminder that 12 months is a convention: in high-ischemic-risk patients who had completed 12 event-free months of DAPT, continuing to 24 months met noninferiority for net adverse clinical events but was associated with markedly fewer ischemic events than clopidogrel monotherapy (composite hazard ratio 2.33, 95% CI 1.47–3.69, for monotherapy),^17^ and in DAPT-MVD, extending DAPT beyond 12 months in patients with multivessel disease reduced ischemic events compared with aspirin alone.^18^ These trials lie beyond our exposure axis, but they frame our result: the shortest duration at which excess risk can be excluded is shortest relative to a reference that may itself be too short for some patients.

These findings have three clinical implications. First, durations under 2 weeks cannot be regarded as safe under any model; both trials that withdrew aspirin at the index procedure failed noninferiority.^24,25^ Second, at 3 months, tested by five units and the position where guideline recommendations and consensus statements for P2Y12 inhibitor monotherapy now sit,^1–4^ the odds ratio for myocardial infarction is 1.09 (95% CrI 0.98–1.25) and major bleeding is reduced by 38%. Third, whether an upper credible limit of 1.25 at 3 months is acceptable is a judgment about the patient, not a property of the evidence: for a patient at high bleeding risk the bleeding curve dominates, for one at high ischemic risk the upper limit does. The curve gives that judgment a scale; it does not remove it. The 158-day estimand is not a recommended duration. It is where a strict bound is met today under the primary function, and 282 days under the alternative; both will shorten as trials at the short end accumulate: the three day-0 trials contributed 5,825 of 58,068 patients.

### Study limitations

As an aggregate-data analysis, this study cannot examine how events accrue in the days after aspirin is withdrawn, the natural next study with individual patient data; it cannot separate duration from agent, landmark design, region, or clinical presentation within the 30-day node, because no trial in the clopidogrel, landmark, discretionary-agent, or mixed-presentation strata contributed a day-0 arm, and no curve specific to chronic coronary syndrome can be estimated; and it treats each trial’s 12-month cumulative count as the outcome, with study baselines absorbing observation-window differences. The axis is sparse between 90 and 365 days, where a single trial at 180 days contributes; the location of the credible bound therefore depends on the fitted function (158 days with Emax, 282 days with the spline), and the primary analysis assumes a single dose-response curve across trials that differ in clinical presentation, P2Y12 inhibitor, and landmark randomization, an assumption that the analyses excluding landmark units (187 days) and restricted to acute coronary syndrome trials (238 days) show to be an approximation. The estimand has Monte Carlo error comparable to the 7-day agreement rule we prespecified; despite tripled chains, the result should not be read to the day. Two reporting rules, for a non-monotone bound and for functions of nearly equal fit, and two sensitivity analyses (S9 and S10) were adopted after the first results were available (Supplementary Table S4). Achieved duration was unavailable for two trials and approximate for one, and removal of single trials moved the estimand between 133 and 179 days. Most trials at the short end of the axis enrolled East Asian patients and the two non-East-Asian day-0 trials were small; extrapolation to populations with a different balance of ischemic and bleeding risk should be cautious.^4,10^

### CONCLUSIONS

Across 16 randomized trials, shortening DAPT before aspirin withdrawal continuously reduced major bleeding, and the ischemic cost of withdrawal declined with the duration of preceding DAPT; a step at 1 month was not supported. A 15% excess of myocardial infarction relative to 12-month DAPT could be excluded from about 5 months with the primary function and about 9 months with the alternative; at 5 months major bleeding was still reduced by 26%. By expressing both risks as continuous functions of duration, these curves allow clinicians and patients to choose the duration of DAPT according to the ischemic margin they accept, rather than a fixed time point.

## Data Availability

The extracted arm-level data, analysis code, and model outputs that support the findings of this study are deposited at Zenodo (https://doi.org/10.5281/zenodo.23250940) and will be made publicly available at the time of publication. The study used aggregate data from published trials and author-supplied aggregate counts; no individual participant data were used.

https://www.crd.york.ac.uk/PROSPERO/view/CRD420261480692

https://doi.org/10.5281/zenodo.23250940

## Author Contributions

J.-S.J. conceived and designed the study, performed the searches, screening, data extraction, and statistical analysis, and drafted the manuscript; K.-H.K. adjudicated screening disagreements, performed the blinded re-screen, and revised the manuscript; J.S.P. performed independent data extraction and risk-of-bias assessment, contributed to the certainty ratings, and revised the manuscript; E.-M.L. and M.-Y.R. contributed to the interpretation of the findings and critically revised the manuscript. All authors approved the final version. J.-S.J. is the guarantor.

## Acknowledgments

The authors thank Hirotoshi Watanabe (Hirakata Kohsai Hospital, Japan) and Takeshi Kimura (Kyoto University Graduate School of Medicine, Japan) (STOPDAPT-2 and STOPDAPT-2 ACS), Jeehoon Kang and Hyo-Soo Kim (Seoul National University Hospital, Republic of Korea; HOST-BR), and Ki Hong Choi and Joo-Yong Hahn (Samsung Medical Center, Sungkyunkwan University School of Medicine, Republic of Korea; SMART-CHOICE) for providing unpublished data from their trials. Generative artificial intelligence (Claude, Anthropic) was used under the authors’ direction to draft analysis code, manage data files, and edit language; all analyses, interpretations, and text were verified by the authors, who take full responsibility for the content.

## Non-standard Abbreviations and Acronyms

BARC: Bleeding Academic Research Consortium
CrI: credible interval
CI: confidence interval
DAPT: dual antiplatelet therapy
DIC: deviance information criterion
GRADE: Grading of Recommendations Assessment, Development and Evaluation
MBNMA: model-based network meta-analysis
OR: odds ratio
PCI: percutaneous coronary intervention. Trial acronyms are expanded in the footnote to Table 1.

## Supplemental Material

Supplemental Methods

Appendix S1

Tables S2–S8 Figures S1–S6

PRISMA 2020 checklist

---

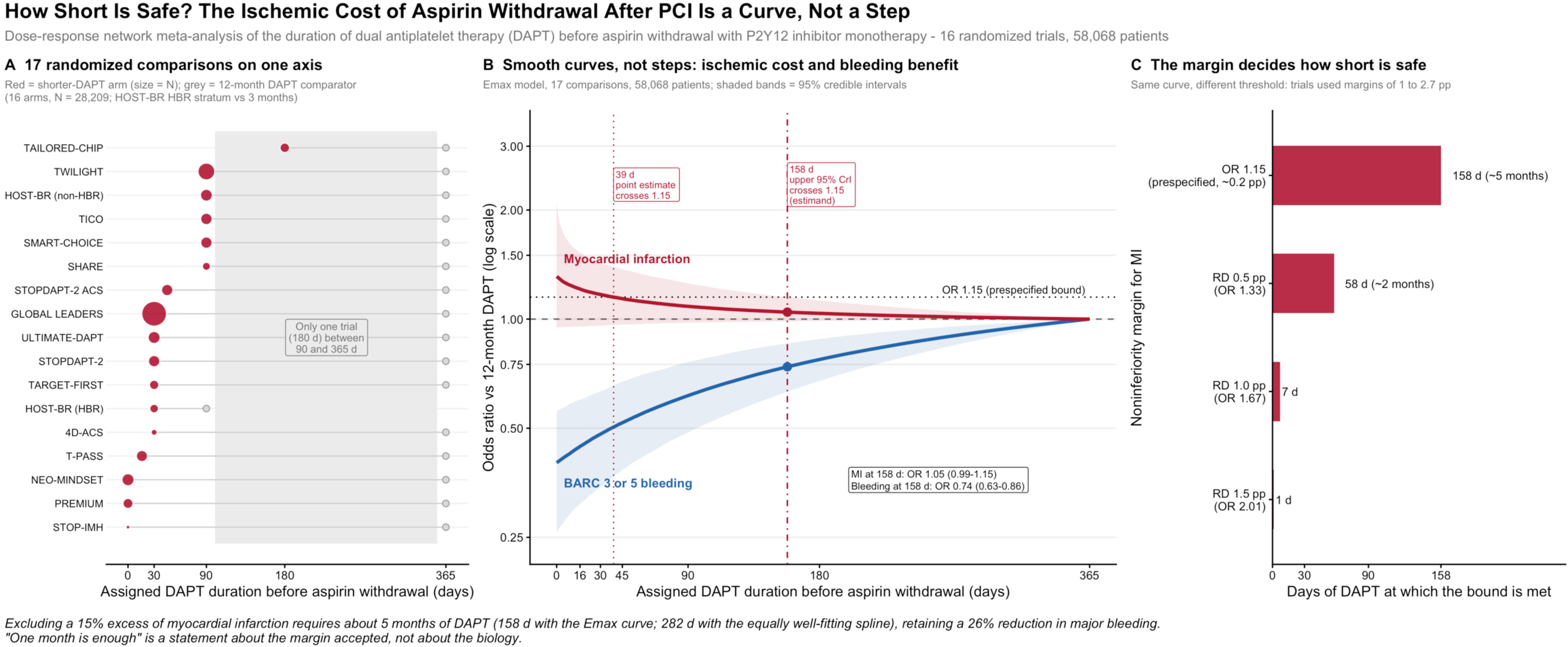

